# Physical activity and *APOE* neuropathology score modify the association of age and [^11^C]-PiB-PET amyloid burden in a cohort enriched with risk for Alzheimer’s disease

**DOI:** 10.1101/2025.03.01.25323157

**Authors:** Eli G. Blum, Kyle J. Edmunds, Brianne Breidenbach, Noah Cook, Ira Driscoll, Sarah R. Lose, Barbara B. Bendlin, Yue Ma, Bradley Christian, Tobey J. Betthauser, Mark Sager, Sanjay Asthana, Sterling C. Johnson, Dane B. Cook, Ozioma C. Okonkwo

**Author notes:** Corresponding Authors: Kyle J. Edmunds, Reykjavík University, Menntavegur 1, 102 Reykjavík, Iceland, Ozioma C. Okonkwo, Clinical Science Center, 600 Highland Ave, J5/156m, Madison, WI 53792.

## Abstract

**Background:** Physical activity (PA) is a protective factor against amyloid-β (Aβ) accumulation in adults at risk for Alzheimer’s disease (AD). This association, however, may differ by apolipoprotein E (*APOE*) genotype. This work examines interactions between age, PA, and neuropathology-based genetic risk for AD (*APOE*_*np*_) on Aβ burden in cortical regions sensitive to its accumulation.

**Materials and Methods:** Included were 388 cognitively unimpaired, older (mean age ± SD = 68.10 ± 7.09; 66% female) participants from the Wisconsin Registry for Alzheimer’s Prevention (WRAP) study. The cohort was enriched with both family history of AD at enrollment and a higher overall prevalence of APOE ε*4* allele carriage than typically observed in the general population. PA was assessed using a self-reported questionnaire. Aβ burden was measured using Pittsburg Compound B (^11^C-PiB) PET imaging, which allowed us to derive volume corrected distribution volume ratio (DVR) maps from nine bilateral regions of interest (ROIs) and a global cortical composite score. Linear regression models examined the interactions between age, PA, and *APOE*_*np*_ on Aβ burden. Finally, *APOE*_*np*_ scores were aggregated according to estimated risk to illustrate the differential effects between active (weekly moderate PA ≥ 150 minutes) and inactive individuals.

**Results:** Three-way interactions (Age × PA × *APOE*_*np*_) were significant (all *P*’s ≤ 0.05) for the global cortical composite and six of the examined ROIs (the PPC, ACC, mOFC, SMG, MTG, and STG). Models stratified by *APOE*_*np*_ and PA showed greater levels of age-related Aβ accumulation in each of these ROIs, with the greatest effects in inactive participants with high *APOE*_*np*_ scores.

**Conclusion:** Individuals with high *APOE_np_* scores who concomitantly engage in suboptimal weekly moderate-intensity PA have greater Aβ burden. These findings underscore how both PA and *APOE*_*np*_ haplotype play intersect in modifying age-related Aβ burden in brain regions susceptible to its deposition in cognitively unimpaired, older adults at risk for AD.

## Introduction

The measurement of fluid and neuroimaging biomarkers has become central to characterizing preclinical Alzheimer’s disease (AD). As one of the core pathophysiological features of AD, amyloid-β (Aβ) deposition in the brain—or amyloidosis—is detectable decades before the onset of clinical symptoms^1^. Aβ is produced when the amyloid precursor protein (APP) is cleaved by beta- and gamma-secretases, generating peptides of varying lengths (38, 40, or 42 amino acids). The 42-amino acid variant is particularly toxic, driving Aβ plaque aggregation—one of the canonical mechanims of AD pathogenesis^2^.

Among the established risk factors for the disease, the apolipoprotein E (*APOE*) gene is one of the most studied. *APOE* is a key regulator of lipid metabolism and cholesterol transport, with its three primary alleles—ε*2*, ε*3*, and ε*4*—determined by two single nucleotide polymorphisms, rs429358 and rs7412, where the ε*2* allele is associated with a TT, ε*3* with TC, and ε*4* with CC haplotype^3^. Each of these alleles confers distinct risk for AD and its biomarker profiles; the ε*3* allele serves as a neutral reference, the ε*4* allele is strongly associated with increased amyloidosis and elevated AD risk, whereby carrying one ε*4* allele increases AD risk three- to four-fold and carrying two ε*4* alleles increases risk up to twelvefold^4^. In contrast, the ε*2* allele is associated with lesser Aβ plaque aggregation and a 50% lower risk for AD^5,6^. Given these opposing effects, AD biomarker studies should account for the differential influence of separate *APOE* haplotypes to provide a more nuanced understanding of genotype-specific interactions.

Physical activity (PA) has emerged as a potential modulator of both age-related Aβ burden^7^ and AD risk^8^—particularly among ε*4* carriers^9^. An evolving body of work suggests that PA may attenuate high Aβ levels in AD-vulnerable cortical regions^10^ and that an inverse relationship between PA and plasma Aβ levels may reflect improved peripheral clearance of Aβ in physically active individuals^11^. Recent findings from our group^7,12,13^ and others^14^ suggest that PA attenuates age-related increases in core AD pathology; mechanistically, greater engagement in PA is associated with increased cerebral blood volume, which supports neuronal survival, neurogenesis, and cognitive function^15,16^. However, the precise mechanisms underlying the relationship between PA and Aβ aggregation or other AD-related pathophysiological processes remains an area of active investigation—especially within the context of *APOE* risk.

The present study aims to examine whether genotype-specific neuropathology-based risk for AD (*APOE*_*np*_) and PA modify the relationship between age and both global and regional cortical ^11^C-PiB PET binding across nine bilateral brain regions of interest (ROIs) known to be susceptible to AD-related amyloidosis^17–19^ in a cognitively unimpaired, middle-aged and older cohort enriched for AD risk.

## Materials and Methods

### Participants

Participants in this study (n=388; mean age ± SD = 68.10 ± 7.09; 66% female) were selected based on data availability from the Wisconsin Registry for Alzheimer’s Prevention (WRAP) study, which consists of approximately 1700 cognitively unimpaired late to middle-aged adults who were between the ages 40 and 65 at study entry^20^. All participants completed a PA questionnaire, and a PET brain scan. The Institutional Review Board of the University of Wisconsin gave ethical approval for this work and approved all study procedures. All study participants provided signed informed consent before participation.

### Physical activity assessment

PA was quantified by calculating the average weekly total minutes of moderate-intensity activity reported by each participant on the Women’s Health Initiative physical activities questionnaire^7,21^. This questionnaire inquired about current frequency and duration of various physical activities, including walking outside the home for >10 minutes and engagement in mild (e.g., slow dancing, golf), moderate (e.g., calisthenics, easy swimming), or vigorous (e.g., jogging, aerobics) exercise. For moderate-level activities, the reported frequencies ranged from rarely to ≥7 times/week and durations ranged from 15 minutes to 1 hour/session. Finally, the total weekly duration of moderate-level PA was calculated by multiplying participants’ weekly frequency by their average duration of each session; this final estimate ranged from 15 to 360 minutes of weekly moderate PA within the cohort. Finally, to illustrate the three-way interaction of Age×PA×*APOE*_*np*_, models were stratified by both *APOE*_*np*_ and PA such that ‘Active’ and ‘Inactive’ participants were defined by the achievement of ≥150 versus < 150 weekly minutes of moderate-intensity PA, as recommended by the US Department of Health and Human Services (DHHS) Physical Activity Guideline^22^.

### Positron emission tomography (PET) imaging protocol

^11^C-PiB PET imaging for this study was performed as previously described^23^. Briefly, PET data were acquired using a 70-minute dynamic acquisition followed by reconstruction with a filtered back-projection algorithm. Data were corrected for random events, attenuation of annihilation radiation, dead time, scanner normalization, and scatter radiation. Realignment and coregistration to T1 MRI images was performed using SPM 8 (www.fil.ion.ucl.ac.uk/spm). Finally, PET images were transformed into voxel-wise distribution volume ratio (DVR) maps of PiB binding using the Logan method, with cerebellar gray matter as the reference region.

Using the Automated Anatomical Labeling atlas implemented in the WFU PickAtlas toolbox^24^, a series of binary masks was employed to sample ^11^C-PiB-PET uptake from nine regions of interest (ROIs) that are susceptible to early Aβ aggregation^18,25–27^: anterior cingulate gyrus (ACC), angular gyrus (ANG), posterior cingulate gyrus (PCC), superior and inferior parietal lobules (posterior parietal cortex, PPC), medial orbitofrontal cortex (mOFC), middle temporal gyrus (MTG), precuneus (PREC), supramarginal gyrus (SMG), and superior temporal gyrus (STG). Left and right DVR values for each ROI were averaged and then normalized to their respective regional volumes (in cm^3^)^28^. Values from all nine ROIs were also aggregated into a global cortical composite score. Further details on ^11^C-PiB radiotracer synthesis, PET and MRI data acquisition and processing, and DVR map generation have been previously published^7,23^.

### *APOE* _***np***_ risk assessment

Participants in this study were previously genotyped using competitive allele-specific PCR-based KASP^29^. A log-transformed index of neuropathology-based genetic risk for AD (*APOE*_*np*_) was assigned to each participant as previously described^30^ to incorporate both the risk associated with ε*4* allele carriage and the protective effects of carrying the ε*2* allele. For this measure, a negative overall score indicates a protective effect, while a positive score reflects increased risk for AD, with a score of zero corresponding to the reference risk of ε*3*ε*3* carriage: the total distribution of *APOE*_*np*_ scores was: n=1 for ε*2*ε*2* (*APOE*_*np*_ = −1.833), n=34 for ε*2*ε*3* (*APOE*_*np*_ = −0.916), n=205 for ε*3*ε*3* (*APOE*_*np*_ = 0), n=9 for ε*2*ε*4* (*APOE*_*np*_ = 0.904), n=114 for ε*3*ε*4* (*APOE*_*np*_ = 1.742), and n=25 for ε*4*ε*4* (*APOE*_*np*_ = 3.293).

### Statistical analyses

Cross-sectional linear regression models were employed to test three-way interactions between age, PA, and *APOE*_*np*_ on global and regional ^11^C-PiB binding. All models covaried for sex, parental family history of AD, and age difference between the time of PA survey and ^11^C-PiB PET imaging. All analyses were completed with RStudio version 4.3.2; findings with p<0.05 were considered significant.

## Results

As shown in Table 1, the current sample consisted of n=388 participants, of whom 35 (9%) had a negative (protective) *APOE*_*np*_ score, 205 (53%) had a reference *APOE*_*np*_ score of 0, and 148 (38%) had a positive *APOE*_*np*_ score (increased risk for AD). The average age of participants at the time of their PET scan was 68.10 ± 7.09 years, and 66% were female with 95.9% White, 1.8% Black or African American, 1.5% American Indian or Alaska Native, and 0.3% Asian; race data was unavailable for 0.5% of the cohort. Mean volume corrected ^11^C-PiB PET DVR scores were highest in the increased risk group (*APOE*_*np*_ > 0) and lowest in the protected group (*APOE*_*np*_ < 0) for both the global composite and each of the ROIs. Mean weekly minutes of moderate-level PA was likewise highest in the increased risk group and lowest in the protected group.

**Table 1.** Participant characteristics.

| *Variable:* | *APOE<sub>np</sub> score* |  |  |
| --- | --- | --- | --- |
| | **Protected**<br>($APOE_{np} < 0$) | **Reference**<br>($APOE_{np} = 0$) | **Increased Risk**<br>($APOE_{np} > 0$) |
| *n* | 35 | 205 | 148 |
| Age (years), mean (SD) | 69.2 (8.03) | 68.8 (7.23) | 66.9 (6.46) |
| Age difference (years), mean (SD) | 3.88 (4.20) | 3.05 (3.24) | 3.03 (3.01) |
| Women, *n* (%) | 24 (68.6%) | 132 (64.4%) | 99 (66.9%) |
| Race, *n* (%) |  |  |  |
| Black or African American | 1 (2.9%) | 3 (1.5%) | 3 (2.0%) |
| American Indian or Alaska Native | 0 (0%) | 5 (2.4%) | 1 (0.7%) |
| Asian | 0 (0%) | 1 (0.5%) | 0 (0%) |
| White | 34 (97.1%) | 194 (94.6%) | 144 (97.3%) |
| Unavailable | 0 (0%) | 2 (0.98%) | 0 (0%) |
| *APOE* genotype, *n* (%) |  |  |  |
| $\epsilon 2\epsilon 2$ ($APOE_{np} = -1.833$) | 1 (2.9%) | | |
| $\epsilon 2\epsilon 3$ ($APOE_{np} = -0.916$) | 34 (97.1%) | | |
| $\epsilon 3\epsilon 3$ ($APOE_{np} = 0$) | | 205 (100.0%) | |
| $\epsilon 2\epsilon 4$ ($APOE_{np} = 0.904$) | | | 9 (6.1%) |
| $\epsilon 3\epsilon 4$ ($APOE_{np} = 1.742$) | | | 114 (77.0%) |
| $\epsilon 4\epsilon 4$ ($APOE_{np} = 3.293$) | | | 25 (16.9%) |
| Weekly moderate-level PA (mins), mean (SD) | 88.7 (75.0) | 104.4 (87.0) | 111.4 (82.1) |
| Weekly moderate-level PA $\geq 150$ minutes, *n* (%) | 13 (37.14%) | 121 (58.5%) | 96 (63.6%) |
| $^{11}\text{C}$-PiB-PET DVR (volume corrected), mean (SD) | | | |
| *Global composite* | 0.274 (0.025) | 0.285 (0.053) | 0.330 (0.083) |
| ACC | 0.228 (0.028) | 0.228 (0.042) | 0.270 (0.074) |
| ANG | 0.204 (0.022) | 0.212 (0.044) | 0.247 (0.068) |
| mOFC | 0.367 (0.041) | 0.379 (0.072) | 0.450 (0.131) |
| MTG | 0.060 (0.006) | 0.061 (0.011) | 0.069 (0.017) |
| PCC | 1.112 (0.135) | 1.175 (0.246) | 1.349 (0.358) |
| PPC | 0.085 (0.010) | 0.091 (0.024) | 0.106 (0.030) |
| PREC | 0.100 (0.010) | 0.106 (0.024) | 0.125 (0.035) |
| SMG | 0.189 (0.018) | 0.190 (0.033) | 0.215 (0.050) |
| STG | 0.122 (0.014) | 0.123 (0.021) | 0.137 (0.032) |

*Abbreviations:* ACC = anterior cingulate; ANG = the angular gyrus; $APOE_{np}$ = $APOE$ neuropathology risk score; $^{11}\text{C}$ -PiB-PET = Pittsburgh Compound-B positron emission tomography; DVR = distribution volume ratio; mOFC = medial orbitofrontal cortex; MTG = middle temporal gyrus; PA = average weekly total minutes of moderate level exercise; PCC = posterior cingulate; PPC = posterior parietal cortex; PREC = precuneus; SMG = supramarginal gyrus; STG = superior temporal gyrus.

Table 2 present results from moderation models that examined whether associations between age and ^11^C-PiB binding were modified by PA and *APOE*_*np*_ score. The three-way Age×PA×*APOE*_*np*_ interaction was significant for the global cortical composite. Additionally, the three-way interaction was significant for six of the nine ROIs examined: the ACC, MTG, mOFC, PPC, SMG, and the STG. In addition to the three-way interaction (Age×PA×*APOE*_*np*_), Table 2 also contains the results of the main effects and all two-way interactions (Age×PA, Age×*APOE*_*np*_, PA×*APOE*_*np*_) from the same models.

**Table 2.** Results from global cortical composite and regional ^11^C-PiB PET moderation models.

|  | *Global Composite* |  | ACC |  | ANG |  | mOFC |  | MTG |  | PCC |  | PPC |  | PREC |  | SMG |  | STG |  |
| --- | --- | --- | --- | --- | --- | --- | --- | --- | --- | --- | --- | --- | --- | --- | --- | --- | --- | --- | --- | --- |
| *Variable* | **B**<br>**(SE)** | **p** | **B**<br>**(SE)** | **p** | **B**<br>**(SE)** | **p** | **B**<br>**(SE)** | **p** | **B**<br>**(SE)** | **p** | **B**<br>**(SE)** | **p** | **B**<br>**(SE)** | **p** | **B**<br>**(SE)** | **p** | **B**<br>**(SE)** | **p** | **B**<br>**(SE)** | **p** |
| **Age** | 0.049<br>(0.011) | **<0.001** | 0.049<br>(0.013) | **<0.001** | 0.047<br>(0.011) | **<0.001** | 0.030<br>(0.005) | **<0.001** | 0.042<br>(0.010) | **<0.001** | 0.059<br>(0.014) | **<0.001** | 0.052<br>(0.013) | **<0.001** | 0.063<br>(0.014) | **<0.001** | 0.037<br>(0.010) | **<0.001** | 0.034<br>(0.010) | **<0.001** |
| **PA** | -0.020<br>(0.012) | 0.086 | -0.022<br>(0.013) | 0.101 | -0.014<br>(0.012) | 0.237 | -0.008<br>(0.005) | 0.118 | -0.015<br>(0.010) | 0.158 | -0.023<br>(0.014) | 0.264 | -0.026<br>(0.013) | **0.044** | -0.027<br>(0.015) | 0.059 | -0.013<br>(0.010) | 0.198 | -0.015<br>(0.001) | 0.144 |
| **APOE<sub>np</sub>** | 0.087<br>(0.010) | **<0.001** | 0.101<br>(0.011) | **<0.001** | 0.081<br>(0.01) | **<0.001** | 0.038<br>(0.004) | **<0.001** | 0.069<br>(0.009) | **<0.001** | 0.101<br>(0.012) | **<0.001** | 0.090<br>(0.011) | **<0.001** | 0.106<br>(0.012) | **<0.001** | 0.062<br>(0.009) | **<0.001** | 0.064<br>(0.008) | **<0.001** |
| **Age × PA** | -0.001<br>(0.004) | 0.940 | 0.002<br>(0.013) | 0.910 | -0.003<br>(0.011) | 0.817 | -0.002<br>(0.005) | 0.742 | -0.004<br>(0.010) | 0.667 | 0.005<br>(0.014) | 0.755 | -0.002<br>(0.013) | 0.870 | -0.002<br>(0.014) | 0.917 | -0.004<br>(0.010) | 0.729 | -0.004<br>(0.010) | 0.717 |
| **Age × APOE<sub>np</sub>** | 0.017<br>(0.010) | 0.086 | 0.025<br>(0.011) | **0.029** | 0.013<br>(0.010) | 0.192 | 0.011<br>(0.005) | **0.024** | 0.013<br>(0.009) | 0.162 | 0.020<br>(0.013) | 0.122 | 0.016<br>(0.011) | 0.148 | 0.021<br>(0.013) | 0.103 | 0.010<br>(0.009) | 0.256 | 0.013<br>(0.009) | 0.122 |
| **PA × APOE<sub>np</sub>** | -0.003<br>(0.010) | 0.754 | -0.012<br>(0.012) | 0.321 | -0.005<br>(0.010) | 0.606 | -0.003<br>(0.005) | 0.425 | -0.005<br>(0.009) | 0.593 | 0.005<br>(0.013) | 0.721 | -0.002<br>(0.012) | 0.886 | 0.007<br>(0.013) | 0.621 | -0.006<br>(0.009) | 0.534 | -0.006<br>(0.009) | 0.500 |
| **Age × PA × APOE<sub>np</sub>** | -0.024<br>(0.012) | **0.047** | -0.028<br>(0.014) | **0.044** | -0.023<br>(0.012) | 0.058 | -0.013<br>(0.006) | **0.023** | -0.025<br>(0.011) | **0.019** | -0.016<br>(0.015) | 0.288 | -0.028<br>(0.014) | **0.038** | -0.026<br>(0.015) | 0.084 | -0.024<br>(0.011) | **0.025** | -0.023<br>(0.010) | **0.026** |

*Abbreviations:* ACC = anterior cingulate; ANG = the angular gyrus; APOE<sub>np</sub> = APOE neuropathology risk score; <sup>11</sup>C-PiB-PET = Pittsburgh Compound-B positron emission tomography; DVR = distribution volume ratio; mOFC = medial orbitofrontal cortex; MTG = middle temporal gyrus; PA = average weekly total minutes of moderate level exercise; PCC = posterior cingulate; PPC = posterior parietal cortex; PREC = precuneus; SMG = supramarginal gyrus; STG = superior temporal gyrus.

The interaction effects (Table 2) highlighted several additional findings; notably, the three-way interaction (Age×PA×) was significantly associated with the global cortical composite (β = -0.02; p = 0.05) and DVR scores in six of the nine ROIs, with the most significant effects in the MTG (β= -0.03; p = 0.02) and SMG (β = -0.02; p = 0.03).

To illustrate the three-way interaction effect of Age×PA×, the following three-panel figures were generated: Figure 1 depicts the interaction effects on global cortical ^11^C-PiB burden, while Figure 2 presents three-panel figures for each of the six significant ROIs. In all panels, inactive persons with high had the highest age-related levles of Aβ.

**Figure 1.**
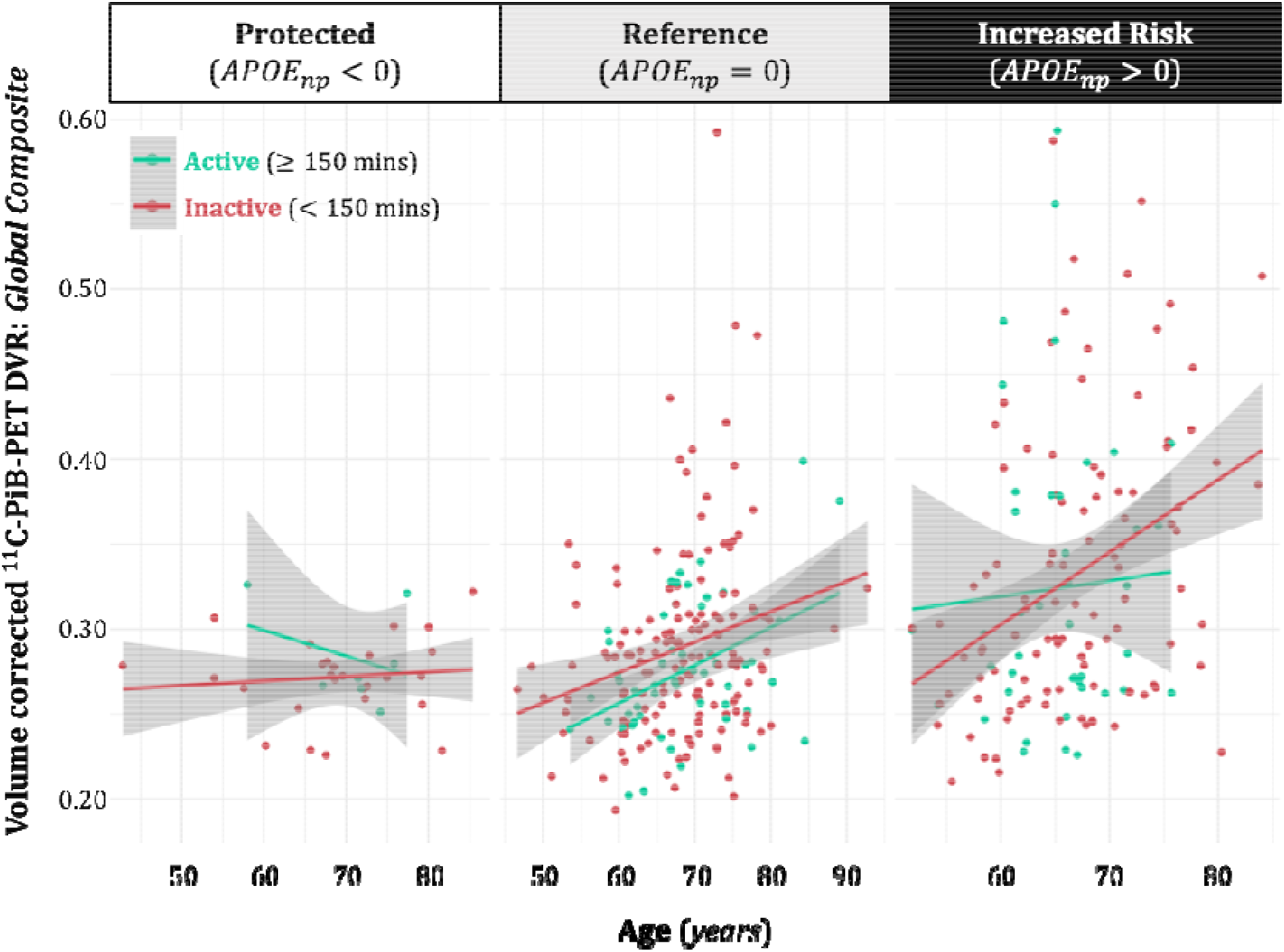
Associations between age and volume corrected global cortical composite ^11^C-PiB PET DVR score, showing differential effects of in Active vs. Inactive (stratified by achievement of the recommended 150 weekly minutes of moderate-level PA).

**Figure 2.**
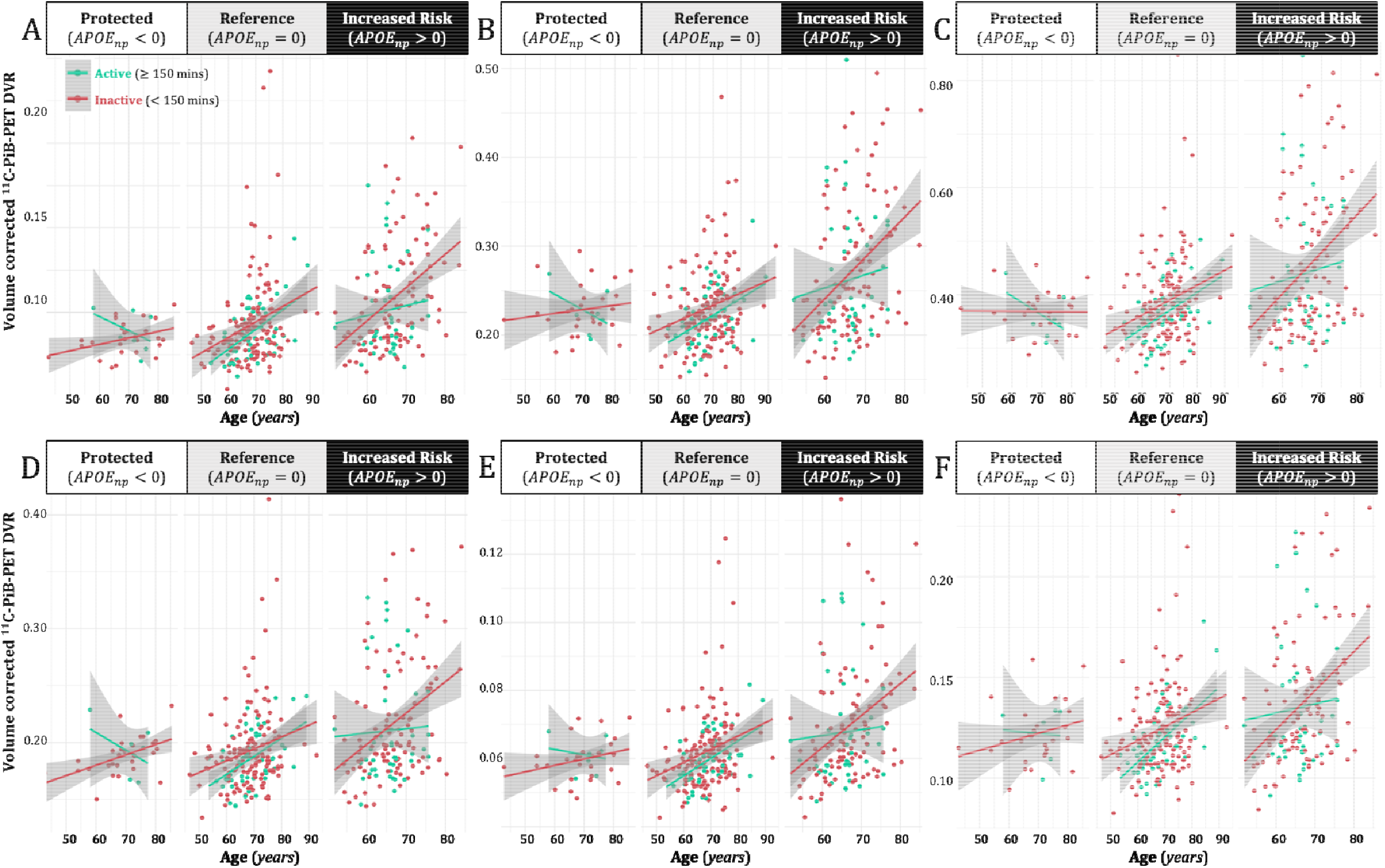
Associations between age and regional volume corrected ^11^C-PiB PET DVR scores. Differential effects of in Active vs. Inactive (stratified by achievement of the recommended 150 weekly minutes of moderate-level PA) in the A) PPC; B) ACC; C) mOFC; D) SMG; E) MTG; and F) STG.

## Discussion

In a cohort enriched with risk for AD, we examined whether genotype-specific, neuropathology-based risk for AD (*APOE*_*np*_) and PA modified the relationship between age and volume corrected ^11^C-PiB PET DVR scores across nine bilateral brain ROIs succeptible to AD-related amyloidosis. Overall, our results suggest that the interactive influence of age, PA, and *APOE*-associated risk on Aβ deposition may vary by cortical region. Specifically, among those in the increased risk *APOE*_*np*_ group, physically active participants had less Aβ burden in the global cortical composite and six of the nine ROIs examined: the PPC, ACC, mOFC, SMG, MTG, and STG.

Our results underscore that the relationship between age and Aβ burden is influenced by both PA and genetic risk based on the *APOE*_*np*_ score. The observed relationship between age and Aβ deposition is well-documented and considered an early marker for the progression to mild cognitive impairment (MCI) or AD^17,31^. The influence of *APOE*_*np*_ on this relationship is visually represented in Figures 1 and 2, where ^11^C-PiB DVR scores were positively associated with both age and *APOE*_*np*_ score in all significant ROIs and the global cortical composite—especially for individuals who did not engage in the recommended 150 minutes of weekly moderate-intensity PA.

As previously mentioned, our group has consistently reported that moderate PA can attenuate deleterious age-related alterations in core AD biomarkers in preclinical AD, including the accumulation of Aβ^7,12,13^. The present study extends these findings by accounting for the differential influence of all *APOE* haplotypes through the *APOE*_*np*_ score. In this regard, the present results suggest that as individuals age, PA may be more beneficial to those with higher *APOE*_*np*_ scores with regards to attenuating Aβ aggregation—particularly in the six regions known to be vulnerable to Aβ deposition in preclinical AD and may therefore represent strategic targets for evaluating the effects of exercise-based interventions.

The presence of at least one ε*4* allele of the *APOE* gene is a well-established risk factor for Aβ accumulation^9^ and the development of AD dementia^8^. Comparatively less is known about how other *APOE* genotypes influence AD biomarkers or how they interact with lifestyle factors such as PA. By utillizing the *APOE*_*np*_ score developed by Deming et al.^30^, the present study accounted for both the protective effects of the ε*2* allele and the risk-enhancing effects of the ε*4* allele. Within the protected group, DVR scores did not significantly increase with age, regardless of PA level. In contrast, age-related changes in PiB PET DVR scores in the reference group were similar for both physically active and inactive individuals—although there was a trend of lower DVR scores in the active group. Finally, in the increased risk group, active individuals had less age-related Aβ accumulation compared to inactive individuals. Together, our findings suggest that PA may play a critical role in moderating Aβ accumulation—particularly in individuals who are genetically predisposed to AD.

The main limitation of the present study is the lack of racial and ethnic diversity in its sample; 96% of the sample identified as white. Expanding this research to include a more diverse population may enhance the generalizability of our findings. Another potential limitation is the reliance upon self-reported PA data, which may be less accurate for measuring absolute intensities of PA than objective measures of exercise or cardiorespiratory fitness^32^— for example, future studies may benefit from incorporating objective fitness measures such as VO_2_ max from graded exercise testing. Finally, the uneven distribution of *APOE* genotypes within our sample may also be seen as a limitation, as it was drawn from a general cross-section of the population. This variability resulted in small subsample sizes for certain genotypes, such as ε*2*ε*2* (n=1) and ε*2*ε*4* (n=9), although this representation in our sample aligns with their respective prevalence in the general population.^30,33,34^ Nevertheless, the comparative rarity of these genotypes limits the statistical power and interpretability of our findings within these groups; as such, future studies may consider intentionally recruiting participants with rare *APOE* genotypes to help extend the generalizability of our—and similar—findings in literature.

## Conclusion

In summary, the current study aimed to provide a more nuanced understanding of whether and how PA and genetic risk for AD neuropathy interact to influence the age-related cortical Aβ burden. Unlike binary measures of ε*4* allele carriage, the *APOE*_*np*_ score accounts for the protective effect of the ε*2* allele, offering a more accurate risk estimate. While our findings align with previous research on the beneficial role of PA in attenuating age-related Aβ accumulation, further research is necessary to fully understand the mechanisms that underly modulation of amyloidosis by PA in the preclinical stages of the AD continuum. The present knowledge gaps nothwithstanding, given the current search for effective ways to prevent, delay, or treat AD, our findings suggest that lifestyle modifications, such as increasing weekly participation in moderate-intensity PA to recommended levels, could provide immediate and accessible strategies for at risk individuals.

## Acknowledgements

This work was supported by National Institute on Aging grants R01 AG062167 (O.C.O.), R01 AG077507 (O.C.O.), R01 AG085592 (O.C.O.), R01 AG027161 (S.C.J.), R01 AG021155 (S.C.J.), R01 AG054059 (C.E.G.) and P30 AG062715 (S.A.); and a Clinical and Translational Science Award (UL1RR025011) to the University of Wisconsin, Madison. This study was supported in part by a core grant to the Waisman Center from the National Institute of Child Health and Human Development (P50 HD105353) and a NIH High-End Instrumentation grant (S10 OD030415). Portions of this research were also supported by the Wisconsin Alumni Research Foundation; and the Veterans Administration, including facilities and resources at the Geriatric Research Education and Clinical Center of the William S. Middleton Memorial Veterans Hospital, Madison, WI. We also thank the staff and study participants of the Wisconsin Alzheimer’s Disease Research Center—without whom this work would not be possible.

## Declarations of interests

All authors have contributed to this work and agree with the presented findings; this manuscript has not been published before, nor is it being considered for publication in another journal. O.C.O. and I.D. are Editorial Board Members of this journal but were not involved in the peer-review process of this article nor had access to any information regarding its peer-review.

## Data availability statement

Request for the data utilized in this study may be directed to the corresponding authors.

## Notes

### Competing Interest Statement

The authors have declared no competing interest.

### Author Declarations

The Institutional Review Board of the University of Wisconsin gave ethical approval for this work and approved all study procedures. All study participants provided signed informed consent before participation.

